# Investigating relations between the NICU speech environment and weight gain in infants born very preterm

**DOI:** 10.1101/2022.05.18.22275280

**Authors:** Komal Kumar, Virginia A. Marchman, Maya Chan Morales, Melissa Scala, Katherine E. Travis

## Abstract

**Background:** Children born very preterm (< 32 weeks gestational age), are at risk for poor growth and adverse neurodevelopmental outcomes. Poor outcomes in preterm children have been attributed to the aversive sounds and relative speech paucity of the neonatal intensive care unit (NICU). Experimental studies that directly expose preterm infants to speech sounds in the NICU find significant improvements in health factors relevant for neurodevelopment. Few studies have examined whether natural variations in the speech environment of the NICU are related to short-term health outcomes in preterm infants. Such data are important for optimizing the sound environment of the NICU.

**Objective:** Examine relations between the NICU speech environment and rate of weight gain during hospitalization, an important determinant of physical health and neurodevelopmental outcomes for preterm infants.

**Methods:** Participants were infants born very preterm (*n* = 20). The speech environment of each infant was assessed at 32-36 weeks postmenstrual age using a speech-counting device known as a Starling. Speech rates were averaged for each infant over the 4-week period. Average rates of weight gain (g/kg/day) were ascertained over the same period. Calories were derived from charted intake (kcals/kg/day). Linear regressions examined associations between weight gain and both caloric intake and speech counts. Control analyses explored whether effects remained after controlling for family visitation, time in incubator, and health acuity.

**Results:** Infants who received more calories gained more weight, accounting for more than 30% of the variance. Importantly, speech counts accounted for nearly 29% additional variance (*p* < .001). These effects were not reduced when controlling for family visitation, time in incubator, or health acuity.

**Conclusions:** Enhancing speech exposure in the NICU may be beneficial for physical growth. NICU infant care plans should consider opportunities to increase speech exposure.

## Introduction

Despite advances in perinatal care practices, neurodevelopmental outcomes in children born preterm have not improved substantially in the past 20 years^1^. Approximately fifty percent of children born very born preterm (<32 weeks gestational age, GA) experience mild to moderate disabilities in multiple cognitive domains such as language, executive functioning and attention^2–4^. Poor neurodevelopmental outcomes in preterm children can have lasting negative effects on children’s future academic and occupational success^4–6^. Optimization of current perinatal care practices is needed to promote positive neurodevelopmental outcomes for children born preterm.

Adverse neurodevelopmental outcomes in children born preterm have been attributed to factors related to the auditory environment of the neonatal intensive care unit (NICU), such as the high levels of noxious sounds (e.g., high-pitched alarms), and the relative paucity of speech ^7–9^. In experimental studies, preterm infants directly exposed to speech sounds in the NICU demonstrate improved health outcomes, including increased oxygen saturation^10,11^, feeding tolerance^12,13^ and decreased apnea and bradycardia events^14^. These factors are also known to be relevant for supporting healthy neurodevelopment. Studies have yet to examine whether natural variations in the speech environment of the NICU are related to short-term health outcomes in preterm infants. Such data are important for clarifying how to optimize the speech environment of the NICU to promote health outcomes in support of neurodevelopment. Such data are also relevant for identifying physical health outcomes that may be sensitive to variations in the NICU sound environment.

In the current study we sought to examine relations between natural variations in the NICU speech environment and rate of weight gain during hospitalization. We focused on weight gain because it is critical for infant health and is associated with neurodevelopmental outcomes during infancy and young adulthood^15–18^. There is also evidence that directed biological maternal sounds, which include filtered speech exposure, are directly related to weight gain velocity^19^. To obtain continuous measurements of the NICU speech environment, we employed a speech counting device, the Starling (Versame, Inc, Menlo Park, CA). We analyzed the NICU speech environment when preterm infants were between 32-36 weeks postmenstrual age (PMA). We hypothesized that infants who experienced higher level of speech exposure at 32-36 weeks PMA, indexed as speech counts from the Starling, would be significantly and positively correlated with greater weight gain during the same developmental period.

## Methods

### Participant Characteristics

A total of 22 infants born preterm were recruited from the NICU at Stanford’s Lucile Packard Children’s Hospital. Inclusion criteria specified preterm birth < 32 weeks gestational age (GA) at birth. All infants were cared for in an open-bay NICU and were deemed stable for intentional sound exposure by evidence-based institutional developmental care protocols. Participants were enrolled between July 2018 and August 2019. Two infants were later excluded due to very short (< 12 hour) periods when estimates of speech environments occurred, yielding a final sample of *n* = 20. Stanford University Institutional Review Board approved the study and informed consent was obtained from all parents/guardians.

### Protocol

Speech counts were derived using automated speech counting technology in a previously commercially-available wearable device called *Starling*. Starling uses speech recognition technology to generate estimates of the amount of vocal activity that is near and clear to the device. The Starling device generates speech counts in real time and does not save an audio-recording, removing concerns about privacy and reducing the potential that the presence of the device may alter speech behavior. Importantly, speech counting algorithms in Starling were designed to be particularly robust in the presence of environmental noise, an advantage in the often-noisy open-bay NICU environment. In a previous study, Starling was found to be sensitive to variations in the amount of speech that occurred in relation to specific NICU care practices.^20^ Moreover, proof-of-concept testing demonstrated that Starling showed substantial correspondence to speech counts obtained from the Language ENvironment Analysis (LENA) digital language processor (LENA Research Foundation, Boulder, CO), a speech recording device that has been used in studies assessing NICU sound environments^9^.

Parents or guardians of potential participants were approached by a research assistant as soon as possible after the infant’s birth. After obtaining informed consent, two Starling devices were placed at the head of each infant’s bed (either incubator or open crib) to allow us to obtain continuous round-the-clock estimates of the infant’s speech environment from time of study enrollment until hospital discharge. Approximately every 5 days, a research assistant would recharge, sync, and download daily speech counts per 5 minutes from cloud-based servers. The positions of the Starling devices were also checked and timing of changes in bed type were noted (e.g., the infant moved from an incubator to an open crib). Later, a research assistant checked the speech count outputs for quality, identifying and removing segments in which counts were inconsistent across the two devices or if there was evidence that one or both of the devices was malfunctioning.

### Measures

#### Demographics and Clinical Variables

At or near time of enrollment, parents or guardians completed two questionnaires from which demographic characteristics, including sex, race/ethnicity, and language background, were derived. Socioeconomic status (SES) was estimated using a modified version of the Hollingshead Index^21^, which takes into account both parents’ education and occupational levels. To characterize medical conditions, we extracted the following information from participants’ electronic medical record: presence of medical or surgical necrotizing enterocolitis (NEC), bronchopulmonary dysplasia (BPD, defined as treatment with supplemental oxygen at 36 weeks PMA), severe intraventricular hemorrhage (IVH, defined as presence of IVH grade 3 or higher, and sepsis (defined as a positive blood culture with clinical signs of illness). For analyses, infants were grouped by presence/absence of any of these four conditions, yielding a scale from 0 to 4. Instances of visitation by all family members were charted by clinical staff as part of routine charting. Visits per day was computed for each infant as the number of visitation instances out of number of days in the data collection period.

#### Speech Counts

Given variable lengths in data collection due to factors such as differences in GA at birth and/or length of hospital stay, we approximated the speech environment of the NICU by analyzing speech counts that occurred within same developmental age period for all infants: 32 weeks; 0 days to 36 weeks; 6 days postmenstrual age (PMA). This age-range was chosen because infants are capable of perceiving human speech sounds at this GA and are more likely to be medically stable and thus able to tolerate speech exposure. Thus, mean counts reported here reflect only speech experiences that occurred during this window, even though some infants may have had data collected prior or after that period. Mean speech/5 min reflects the number of speech units identified by Starling as produced near to the infant, averaged across all 5 minutes segments with available data during the data collection period.

#### Rate of Weight Gain

Average rates of weight gain were computed from infants’ weekly weights (g) measured at 32 weeks; 0 days, 33 weeks; 0 days, 34 weeks; 0 days and 35 weeks; 0 days PMA obtained via medical chart review. To account for differences in weight at start of the period, all weight gain was measured weekly in g/kg/day averaged over the previous 7 days.

#### Caloric intake

Caloric intake (kcal/kg/day) for each infant was measured at two time points: 32 weeks 0 days and 35 weeks 0 days. Total calories were calculated by combining enteral and parental calories, if present. Breast milk was assumed to have 20 kcal/oz and any caloric fortification of feeds was noted. Calories from total parenteral nutrition or intravenous fluids was also gathered, if appropriate. Total caloric intake for a given day (kcal/kg/day) was determined by dividing total daily caloric intake by the weight as measured on the same day. To derive estimates of overall caloric intake across the period, intakes at the two time points were averaged.

#### Statistical Analysis

Descriptive statistics (frequencies, *M, SD*, and range) characterized the demographic and clinical features of study participants, as well as the weight, caloric intake, health acuity, and speech measures. We then conducted a series of linear regression analyses to explore predictors of growth rates, specifically, caloric intake and speech counts. Follow-up analyses controlled for rates of family visitation, time in incubator, and health acuity composite score. Preliminary analyses found that mean speech counts were not normally distributed, and thus, values were log-transformed for analyses. For all analyses, a threshold of *p* ≤ .05 (two sided) was used to determine significance. Exact *p*-values are provided for all significant and non-significant effects when *p* < .001.

## Results

### Demographic and clinical characteristics

Table 1 presents demographic and clinical characteristics of the participants. Participants were infants born at approximately 28 weeks GA, with slightly more males than females. Families reported a mix of racial backgrounds, with about one-quarter of the families reporting Hispanic/Latino ethnicity. Primary language of the maternal caregiver was available for 16 infants, revealing that 44% of the families reported that their primary home language was English. Families from a broad range of socioeconomic backgrounds were represented in the sample.

**Table 1.** Descriptives of the demographic and clinical characteristics of the participants (*n* = 20)

|  | *M (SD)*<br>or *n (%)* | Range |
| --- | --- | --- |
| Sex (% male) | 12 (60) | -- |
| Birth GA<sup>a</sup> | 28.2 (2.8) | 23.0 - 31.7 |
| Race |  |  |
| Asian | 7 (35) | -- |
| African American/Black | 1 (5) | -- |
| Other | 5 (25) | -- |
| White | 7 (35) | -- |
| Hispanic | 5 (25) | -- |
| Primary Language English | 7 (44) | -- |
| Hollingshead SES<sup>b</sup> | 43.2 (20.5) | 8 - 66 |
| Family Visitation<sup>c</sup> | 0.9 (0.3) | 0.4 - 1.7 |
| Health Factors<sup>d</sup> |  |  |
| BPD | 5 (25) | -- |
| NEC | 4 (20) | -- |
| IVH-severe | 5 (25) | -- |
| Sepsis | 1 (5) | -- |
| Health Acuity<sup>e</sup> | 0.7 (1.0) | 0 - 3 |

<sup>a</sup>Gestational Age at birth (weeks)
<sup>b</sup>Socioeconomic status based on a modified version of the Hollingshead four-factor index of socioeconomic status.<sup>21</sup>
<sup>c</sup>Rate of visitation defined as the number of visitation events out of days during study period.
<sup>d</sup>Number of children with BPD (bronchopulmonary dysplasia: treatment with supplemental oxygen at 36 weeks PMA), NEC (necrotizing enterocolitis), IVH (intraventricular hemorrhage of grade 3 or higher), and Sepsis (positive blood culture with clinical signs of illness).
<sup>e</sup>Composite of health conditions (0-4 possible) per infant.

Table 1 also presents the mean rates of visitation during the recording window for the families based on routine charting in the electronic medical record. On average, families visited the NICU just under once per day, although there was considerable variation in visitation rates. Review of health conditions showed that about one-quarter of the infants had each of the four health conditions. A total of 60% of the infants (12 of 20) were reported to not have any of the four conditions and 40% were reported to have one or more conditions (8 of 20). Thus, this sample of infants was relatively healthy, as also indicated by the mean composite score of less than 1.0.

### Speech, Weight, and Caloric Intake Measures

As shown in Table 2, speech counts were derived based on more than 20,000 minutes of data collection per infant, or approximately, 338 hours, and more than 14 days, on average. While lengths of study period were variable across infants, spanning from approximately 12 hours to nearly 24 days, estimates of speech counts were based on at least 7 days for more than 80% of the infants. The study period was about twice as long when infants were in the incubator, as in the open crib. Some infants spent all of the study period in the incubator (*n* = 8), whereas, other infants spent all of the study period in an open crib (*n* = 4). A total of *n*=8 infants spent time in both an incubator and an open crib. Replicating earlier studies^20^, speech counts were higher, on average, when the infants were in open cribs vs. when in incubators. For those infants who were in only one bed type, a between-subjects comparison revealed a significant difference favoring word counts in open cribs vs. incubators, *t*(10) = 2.8, *p* = .02. For those infants who experienced both bed types, a within-subjects paired comparison revealed that infants experienced higher speech counts when in open cribs compared to incubators, *t*(7) = 4.7, *p* = .002. At the same time, speech counts for those infants who experienced both bed types were highly correlated, *r*(7) = .87, *p* = .005, suggesting that regardless of bed type, there was significant variation across infants in the how much speech exposure they experienced. Finally, speech counts, on average, were not significantly different in those families who reported their preferred language as English compared those who reported non-English, *t*(14) = 0.3, *p* = .83.

**Table 2.** Descriptives of length of study period, speech counts, weight, and caloric intake

|  | *M (SD)* | Range |
| --- | --- | --- |
| Days in Study Period |  |  |
| Total | 14.1 (7.1) | 0.5 - 24.2 |
| Incubator | 9.4 (7.5) | 0 – 22.9 |
| Open Crib | 4.7 (4.6) | 0 – 13.9 |
| Minutes in Study Period |  |  |
| Total | 20,287.5 (10,161.7) | 745-34,830 |
| Incubator | 13,598.3 (10,843.5) | 0-32,915 |
| Open Crib | 6,709.3 (6,564.5) | 0-20,085 |
| Speech/5 mins |  |  |
| Total | 19.0 (9.0) | 8.2-46.6 |
| Incubator | 13.9 (4.6) | 6.6-21.0 |
| Open Crib | 26.7 (11.6) | 8.3-46.6 |
| Weight (g) |  |  |
| Start | 1350.2 (234.1) | 730-1710 |
| End | 1958.4 (350.7) | 1060-2495 |
| Growth (g/kg/day) | 18.9 (3.4) | 12.8-26.2 |
| Calories (kcal/kg/day) | 130.4 (15.0) | 90.1 – 152.6 |

Table 2 also shows that infants weighed about 1300 g at the start of the study period, gaining nearly 600 g by 35 weeks GA. Rates of weight gain were about 19 g/kg/day, although there were individual differences in this measure. On average, infants were receiving about 130 kcals/kg/day, across the measurement period, although again, there was range in these values across infants.

### Relations between Speech Counts and Weight Gain

Table 3 shows multiple regression models exploring prediction to rates of weight gain from caloric intake and speech counts as measured by Starling. Model 1 shows that caloric intake made a significant contribution to weight gains, contributing approximately 31%, *F*(1,18) = 8.2, *p* = .01. Importantly, Model 2 shows that the addition of speech counts contributed an additional 29% of the variance, *F*(1, 17) = 12.4, *p* = .003, nearly doubling the variance accounted for overall, *F*(2, 17) = 12.8, *p* < .001. This effect is illustrated in Figure 1. These results suggest that variation in the amount of speech that infants experience during their NICU stay is associated with overall health improvements as indexed by weight gain, even beyond other important predictors like caloric intake. Finally, Models 3, 4, and 5 show that the association between speech counts and weight gain remained significant after controlling for frequency of family visitation, time in incubator, and health acuity composite score, respectively. None of these factors contributed significant additional variance. Thus, the association between variation in quantity of speech experience and infant weight gain is attributable to variability in the experiences that infants receive in the NICU and are not reducible to whether or not family visited, the type of bed infants experienced, or the overall health of the infant.

**Table 3.** Multiple regression models examining prediction of weight gain (g/kg/day) from caloric intake, speech counts, and visitation (*n* = 20); Unstandardized beta (SE).

|  | Model 1 | Model 2 | Model 3 | Model 4 | Model 5 |
| --- | --- | --- | --- | --- | --- |
| Caloric intake<sup>a</sup> | **0.13 (0.04)\*\*** | **0.12 (0.04)\*\*** | **0.12 (0.04)\*\*** | **0.12 (0.04)\*\*** | **0.12 (0.04)\*\*** |
| Speech count<sup>b</sup> | -- | **9.59 (2.73)\*\*** | **9.74 (2.81)\*\*** | **9.45 (2.84)\*\*** | **9.36 (2.72)\*\*** |
| Visitation<sup>c</sup> | -- | -- | -0.70 (1.62) | -- | -- |
| Incubator (mins)<sup>d</sup> | -- | -- | -- | -0.01 (0.01) | -- |
| Health Acuity<sup>e</sup> | -- | -- | -- | -- | -0.56 (0.50) |
| $r^2$-change | **31.2%\*\*** | **28.9%\*\*** | 0.4% | 0.2% | 2.9% |
| $R^2$ | -- | **60.2%\*\*** | **60.6%\*\*** | **60.4%\*\*** | **63.0%\*\*\*** |

Note: \* $p < .05$ , \*\* $p < .01$ , \*\*\* $p < .001$
<sup>a</sup>kcal/kg/day indexed as the total number of calories from enteral and parenteral sources divided by weight in kilograms averaged over measurements taken at 32 weeks; 0 days and 35 weeks; 0 days.
<sup>b</sup>Average number of speech units per 5 mins over the study period
<sup>c</sup>Number of instances of visitation out of days in study period
<sup>d</sup>Number of minutes in incubator across study period
<sup>e</sup>Health acuity composite score as measured by presence/absence of 4 conditions associated with prematurity (i.e., NEC, IVH, BPD and sepsis).

**Figure 1.**
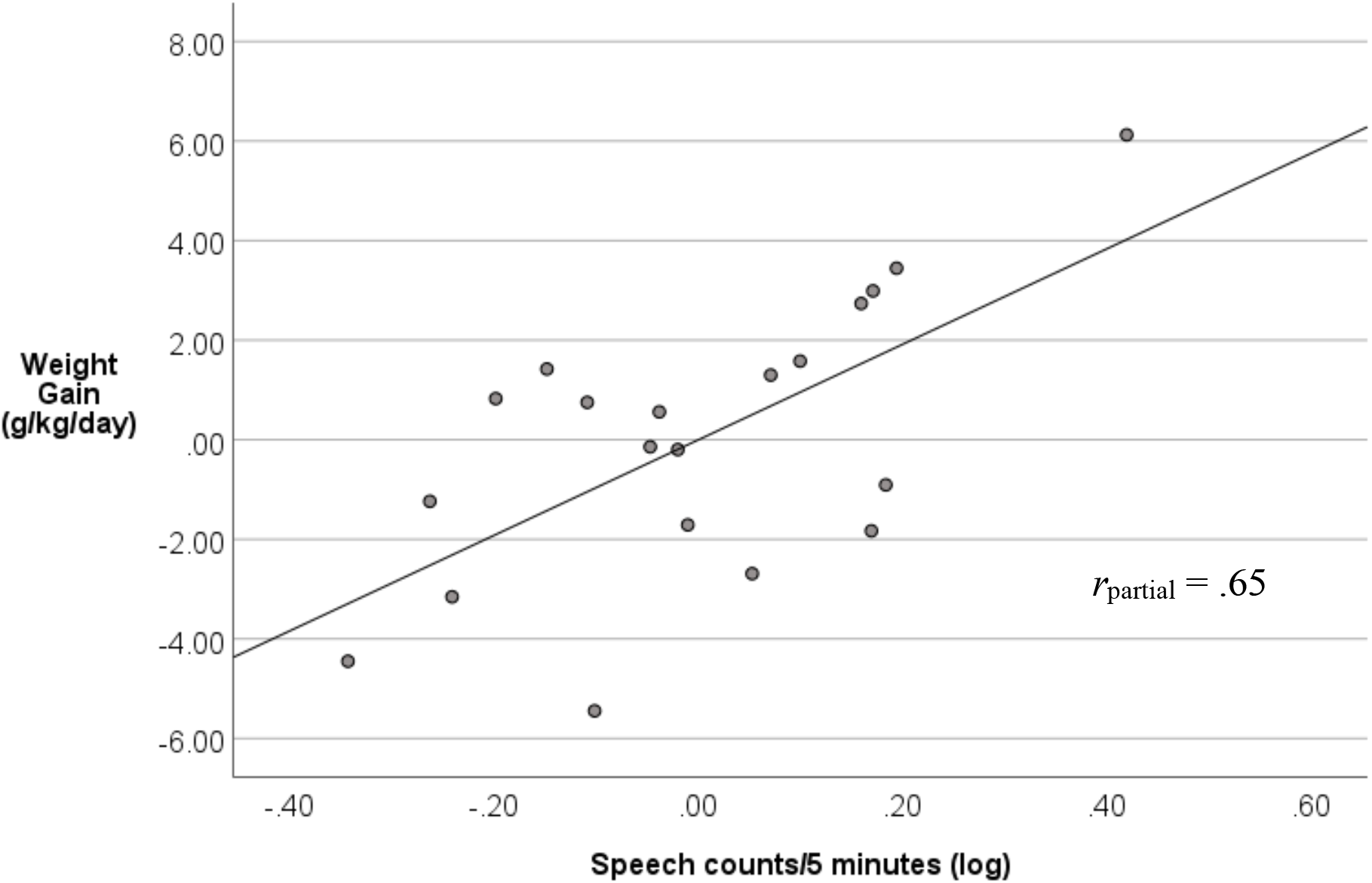
Association between speech counts and weight gain, controlling for caloric intake at start of study period (*n* = 20).

## Discussion

In this study, we documented positive associations between preterm infant weight gain and amounts of speech infants experienced in the NICU. Importantly, speech counts were found to contribute significantly to variance in infants’ weight gain beyond that accounted for by caloric intake, and speech counts were also found to contribute to infants’ weight gain after accounting for family visitation, time in incubator, or health acuity. Together, these finding suggest that the amount of speech that preterm infants are exposed to at bedside may independently account for variations in infants’ physical growth beyond nutritional factors alone. Moreover, other factors which may have influenced speech rates, such as family visitation rates, bed type and infant health, were independent of variation in the amounts of speech experienced by infants from families or clinical staff in the NICU. Overall, these findings contribute to understanding of the NICU speech environment and its potential role in health and neurodevelopmental outcomes in children born preterm.

Previous research has found associations between the NICU speech environment, infant vocalizations and later language outcomes at 7 and 18 months^9,22^. Here, our study provides novel evidence that variations in the NICU speech environment may also play a role in infant weight gain, an important index of infant physical health. Concerns regarding possible stress to infants from sound exposure, and thus worsened weight gain, may impede opportunities for infants to be exposed to speech. In addition, infants with slower weight gain may be kept in an incubator for longer periods, limiting opportunities for speech exposure. Such clinical decisions may be creating or contributing to our measured association. At the same time, the direction of the present association between infant growth and speech exposure merits further discussion. It may have been that infants in our study who gained more weight may have been more active or had more engaging behaviors than infants of lower weights, thus triggering more speech directed to them at bedside. Studies that directly manipulate the amount of speech input provided to preterm infants are likely to be critical for determining whether and how care practices should be modified to enhance speech exposure during NICU hospitalization^23^.

Consistent with our previous work, we found speech counts to be significantly higher for infants in open cribs versus incubators^20^. However, we also found that speech counts were highly correlated for infants that had speech counts measured in both incubators and open cribs. In addition, we also observed that associations between speech counts and infants’ weight gain remained significant after accounting for bed type. Taken together, these findings suggest that the factors that may account for increased speech counts in open-cribs versus incubators (e.g., more overheard speech from staff) are likely to differ from those at are associated with infants’ weight gain. Further studies are needed to identify the specific linguistic factors that may contribute to variability in the NICU speech environment. Such studies could help clarify whether different types of speech (e.g., infant directed speech or “parentese”^24^ versus adult directed speech) and/or sources of speech input (primary caregivers vs clinical staff) are relevant factors that are related to variations in health and later neurodevelopmental outcomes.

Our study window predates the COVID-19 pandemic making evaluation of the impact of any altered care environment impossible. Previous research has shown reduction in parent visitation and in rates of both parent and staff delivered developmental care activities during the COVID-19 pandemic compared to previous periods^25^. Although family visitation did not significantly alter the impact of speech counts on growth in this study, significant reductions in visitation during the pandemic may negatively impact overall opportunities for infant speech exposure. In addition, if staff delivered speech occurs during care times, including developmental care, documented reductions in developmental care during the pandemic may also contribute to reduced speech exposure. Careful examination of growth and neurodevelopment of preterm infants cared for during the pandemic is needed to mitigate possible negative effects.

Limitations of our study include a small number of infants and a variable amount of data available for each infant. Additionally, speech exposure was examined only in the relatively mature window of 32-36 weeks postmenstrual age, so extrapolation of our conclusions regarding the role of speech exposure and weight gain to children at younger gestational ages is impossible. The Starling speech recording device does not allow for listening to the speech environments and thus we are unable to determine the potential sources of variability in speech counts that may have contributed to variations in weight gain. However, the length of our measurements far exceeds that found in previous studies of the speech environment of the NICU making our understanding of the speech environment of our limited number of infants very robust.

Our study adds to the growing body of literature linking speech exposure in the NICU to positive health outcomes. Further research is needed to understand the directionality of this effect. A randomized controlled trial of supplemented speech exposure coupled with speech environment measurement could further explore the value of augmented speech to infant weight gain and would allow for further accounting of possible confounders. As language exposure has been linked to positive neurodevelopmental outcomes, as has optimum growth, the relationship of these factors is another avenue for exploration. As care plans for preterm infants are crafted for optimum outcomes, intentional speech exposure should be attended to as a possible method for intervention.

## Data Availability

All data produced in the present study are available upon reasonable request to the authors

## Conflict of Interest Disclosures

The authors have no conflicts of interest relevant to this article to disclose.

## Notes

**Funding/Support:** This research work was supported by grants from the National Institutes of Health-*Eunice Kennedy Shriver* National Institute of Child Health and Human Development (K.E. Travis, PI; 5R00HD8474904), Stanford Maternal Child Health Research Institute grant (M Scala, PI).

### Competing Interest Statement

The authors have declared no competing interest.

### Funding Statement

This research work was supported by grants from the National Institutes of Health-Eunice Kennedy Shriver National Institute of Child Health and Human Development (K.E. Travis, PI; 5R00HD8474904), Stanford Maternal Child Health Research Institute grant (M Scala, PI).

### Author Declarations

The IRB of Stanford University School of Medicine gave ethical approval for this work.

